# Fractal correlation properties of heart rate variability as a marker of exercise intensity during incremental and constant-speed treadmill running

**DOI:** 10.1101/2023.12.19.23300234

**Authors:** C. R. van Rassel, O. O. Ajayi, K. M. Sales, A. C. Clermont, M. Rummel, M.J. MacInnis

## Abstract

The short-term scaling exponent of detrended fluctuation analysis (DFAα1) applied to interbeat intervals may provide a method to identify ventilatory thresholds and indicate systemic perturbation during prolonged exercise. The purposes of this study were to i) confirm whether DFAα1 values of 0.75 and 0.5 coincide with the gas exchange threshold (GET) and respiratory compensation point (RCP), ii) quantify DFAα1 during constant-speed running near the maximal lactate steady state (MLSS), and iii) assess the repeatability of DFAα1 between MLSS trials. Seventeen runners performed an incremental running test, and eleven and ten runners also performed constant-speed running 5% below, at, and 5% above the MLSS, and a repeat trial at MLSS, respectively. GET (bias [LOA]: –3.6 [–9.1 to 1.9] mL·kg^−1^·min^−1^) and RCP (–3.5 [–14.1 to 7.2] mL·kg^−1^·min^−1^) were overestimated using DFAα1. DFAα1 responses during 30-min running trials near MLSS were variable (i.e., 0.27 to 1.24), and affected by intensity (p=0.019) and duration (p=0.001). No difference in DFAα1 was detected between MLSS trials (p=0.926). These results question whether DFAα1 values can accurately delineate exercise thresholds, but the dependency of DFAα1 on intensity and duration support its potential use to quantify systemic perturbations imposed by continuous exercise.

## Introduction

Training intensity distribution frameworks divide exercise into at least three predetermined intensity zones (Esteve-Lanao et al., 2007; Seiler & Kjerland, 2006). In one model, the moderate, heavy, and severe exercise intensity domains are delineated by the lactate threshold (LT) and maximal metabolic steady state (MMSS), facilitating exercise monitoring and testing (Coates et al., 2023; Jamnick et al., 2020). Methods to identify the LT and MMSS often require multiple invasive laboratory visits, translate poorly to real world settings, and/or incorrectly assume that these physiological attributes are static (Jamnick et al., 2020; Maunder et al., 2021); therefore, simple and translatable methods that consider how these attributes may shift in response to prolonged exercise, previous exercise, or other relevant external and internal stimuli are needed. Such methods may include heart rate variability (HRV) analysis to identify exercise thresholds and monitor physiological shifts from exercise (Gronwald & Hoos, 2020; Kaufmann et al., 2023; Rogers, Giles, Draper, Hoos, et al., 2021; Rogers & Gronwald, 2022)

The short-term scaling exponent alpha-1, derived from detrended fluctuation analysis (DFAα1), is an index that represents fractal, self-similar patterns within a physiological signal like HRV (Goldberger et al., 2002; Peng et al., 1995). To derive DFAα1, the root mean square (RMS) fluctuation of the integrated and detrended HRV RR-interval time-series data is evaluated in observation windows of different sizes (Peng et al., 1995). DFAα1 is then calculated as the slope between the RMS fluctuation data in relation to the different window sizes on a log-log scale (Peng et al., 1995).

During incremental exercise, HRV DFAα1 exhibits an intensity-dependent loss of correlation properties with increasing exercise intensity (Gronwald & Hoos, 2020; Gronwald, Rogers, et al., 2020). At moderate intensities, DFAα1 has values near 1.0, representing correlated patterns. As exercise transitions to the heavy intensity domain, values near 0.75 emerge, indicating a half-way loss of correlation properties. As exercise transitions to the severe intensity domain, values reach 0.5 and lower, indicating uncorrelated or anti-correlated patterns, respectively. This intensity-dependent response has led to suggestions that DFAα1 values of 0.75 and 0.5 represent the first and second exercise thresholds, respectively (Gronwald & Hoos, 2020; Gronwald, Rogers, et al., 2020; Mateo-March et al., 2023; Rogers, Giles, Draper, Hoos, et al., 2021; Rogers, Giles, Draper, et al., 2021a; Schaffarczyk et al., 2023). DFAα1 also responds to exercise training stress. For example, an attenuation of DFAα1 was observed after prolonged ultramarathon running (Rogers, Mourot, et al., 2021) and during a marathon, despite a reduction in running speed (Gronwald et al., 2021). Yet, DFAα1 has not been measured during constant-intensity exercise relative to physiological thresholds, and it is unknown whether DFAα1-derived thresholds determined from incremental exercise translate to constant-intensity exercise.

While DFAα1 is reportedly able to delineate exercise intensity domains, quantify systemic perturbation imposed by a previous bout of exhaustive exercise, and indicate fatigue accumulation during endurance exercise (Gronwald & Hoos, 2020; Rogers & Gronwald, 2022; Rogers, Mourot, et al., 2021; Schaffarczyk et al., 2022), there is limited data to support these suppositions, particularly in running. Accordingly, the purposes of this study were to: i) test whether DFAα1 values of 0.75 and 0.50 coincide with the gas exchange threshold (GET) and respiratory compensation point (RCP), respectively; ii) quantify the DFAα1 response during constant-speed exercise near the maximal lactate steady state (MLSS)—a proxy of the MMSS; and iii) determine the repeatability of DFAα1 at MLSS. We hypothesized that DFAα1 at GET and RCP would not differ from 0.75 and 0.5, respectively; DFAα1 would be stable and ≈ 0.5 at the running speed associated with MLSS; and DFAα1 at MLSS would be repeatable.

## Methods

### Participants

Nineteen (11 male; 8 female) runners (mean [SD]; age = 29.5 [5.2] years; body mass = 68.6 [8.4] kg; height = 172.9 [8.3] cm; ^V̇^ O_2max_, 55.4 [5.6] ml·kg^-1^·min^-1^) classified as recreationally active or trained/developmental (McKay et al., 2022), were recruited using convenience sampling. Written informed consent was provided by the runners prior to participating in the experimental procedures, which were approved by the University of Calgary Conjoint Health Research Ethics Board (REB20-0111 & REB20-1377) and performed in accordance with the Declaration of Helsinki. Prior to test administration, runners completed the Physical Activity Readiness Questionnaire (PAR-Q+) to identify contraindications to exercise testing and ensure participants were free of medical conditions and injuries that could interfere with metabolic and cardiorespiratory exercise responses. All runners provided their own running shoes and wore the same shoes for all testing sessions.

### Experimental Design

All runners visited the laboratory for an initial maximal incremental exercise test (van Rassel et al., 2022). Twelve of these runners (see Table 1 for description) also performed a series of 4 to 5 constant-speed running bouts to determine the running speed 5% below MLSS, at MLSS, and 5% above MLSS, and to perform a repeat trial at the determined speed associated with MLSS (van Rassel et al., 2023). Data were combined from two projects for the incremental exercise analysis; however, seven runners participated in a separate project that did not include constant-speed treadmill running tests and were therefore only included in the incremental analysis. Runners refrained from eating or consuming caffeine within 2 hours prior to their testing sessions and avoided strenuous exercise on the same day prior to the testing sessions. At least 48 hours of time elapsed between exercise sessions. Data from this study were previously published in manuscript that validated a “Step-Ramp-Step” approach to identify the running speed and power associated with the maximal lactate steady state (van Rassel et al., 2022) and a manuscript that investigated the utility of running power as a running performance metric (van Rassel et al., 2023); however, the data presented herein (i.e., DFAα1 data) are distinct and have not been reported elsewhere.

**Table 1.** Incremental exercise testing results, heart rate variability-derived thresholds, and expected DFAα1 for participants that completed incremental and constant-speed testing or incremental testing only.

| | Subject | Sex | $\dot{V}O_{2\max}$ | GET | HRV<sub>GET</sub> | RCP | HRV<sub>RCP</sub> | DFAα1 |
| --- | --- | --- | --- | --- | --- | --- | --- | --- |
|  |  |  | (mL·kg<sup>-1</sup>·min<sup>-1</sup>) | (mL·kg<sup>-1</sup>·min<sup>-1</sup>) | (mL·kg<sup>-1</sup>·min<sup>-1</sup>) | (mL·kg<sup>-1</sup>·min<sup>-1</sup>) | (mL·kg<sup>-1</sup>·min<sup>-1</sup>) | (a.u.)* |
| *Incremental & Constant*<br>*Speed Testing* | 1 | M | 61.7 | 41.4 | 43.2 | 55.0 | 62.1 | 0.59 |
|  | 2 | M | 49.7 | 36.9 | 36.3 | 42.8 | 40.6 | 0.38 |
|  | 3 | M | 60.9 | 37.3 | 40.0 | 52.5 | 48.2 | 0.37 |
|  | 4 | M | 58.2 | 42.0 | 45.9 | 51.1 | 57.8 | 0.64 |
|  | 5 | M | 66.7 | 43.3 | 51.5 | 54.9 | 66.0 | 0.69 |
|  | 6 | F | 53.3 | 36.4 | 42.1 | 44.3 | 45.3 | 0.58 |
|  | 7 | F | 49.5 | 35.1 | 37.7 | 43.5 | 43.5 | 0.50 |
|  | 8 | F | 62.5 | 45.2 | 52.0 | 53.2 | 56.3 | 0.68 |
|  | 9 | F | 51.4 | 36.8 | 40.8 | 46.2 | 44.1 | 0.34 |
|  | 10 | M | 58.2 | 39.9 | 43.2 | 49.5 | 59.8 | 0.66 |
|  | 11 | F | 47.5 | 34.4 | 39.4 | 42.7 | 49.1 | 0.66 |
|  | 12 | F | 48.7 | 33.7 | 38.4 | 42.3 | 43.4 | 0.55 |
| **Average** |  |  | 55.7 | 38.5 | 42.5 | 48.2 | 51.3 | 0.55 |
| **SD** |  |  | 6.5 | 3.7 | 5.1 | 5.0 | 8.6 | 0.13 |
| *Incremental*<br>*Testing Only* | 13 | F | 56.4 | 41.1 | 47.9 | 52.5 | 59.9 | - |
|  | 14 | M | 58 | 38.4 | 39.3 | 49.9 | 45.6 | - |
|  | 15 | M | 54.5 | 40.2 | 40.2 | 50.2 | 53.8 | - |
|  | 16 | M | 56.8 | 41.0 | 40.0 | 47.4 | 61.1 | - |
|  | 17 | F | 55.9 | 34.6 | 41.0 | 47.2 | 47.6 | - |
| **Average** |  |  | 56.3 | 39.1 | 41.7 | 49.4 | 53.6 | 0.55 |
| **SD** |  |  | 1.3 | 2.7 | 3.5 | 2.2 | 7.0 | 0.13 |

\*Expected DFAα1 correspond to the DFAα1 at the respiratory compensation point (RCP) using the linear $\dot{V}O_2$ -DFAα1 regression established during incremental exercise for participants that completed both incremental and constant-speed exercise testing. This DFAα1 value was used to represent the 'expected' DFAα1 response during constant-speed treadmill running tests at the MLSS

### Exercise Protocols

#### Maximal Incremental Exercise Test

Of relevance to the present study, the maximal exercise test involved a moderate intensity step-transition (MOD; 6 min at 1.9 m·s^-1^, 6 min at 2.4 m·s^-1^, and 6 min at 1.9 m·s^-1^) followed by incremental treadmill running (an initial speed of 1.9 m·s^-1^ and increasing by ∼0.2 m·s^-1^ [i.e., 0.5 mph] per min) until volitional exhaustion (van Rassel et al., 2022). Incremental tests were considered maximal if a plateau in ^V̇^ O_2_ was observed (i.e., < 150 mL·min^-1^) despite an increase in running intensity, or if subjects HR was ± 10 bpm of their age-predicted max, RER was greater than 1.15, or [BLa] was at least 8 mmol·L^-1^ upon test completion.

#### Constant Speed Treadmill Running

The MLSS was identified as the highest treadmill speed where the difference between the [BLa] at 10 and 30 min was ≤ 1 mmol·L^-1^ and at least 30 min of exercise was performed (Beneke & von Duvillard, 1996). The constant-speed sessions consisted of 5 min of treadmill running at 1.9 m·s^-1^, followed by treadmill running at the pre-determined testing speed (van Rassel et al., 2022). Constant-speed sessions were performed until running tests were completed at speeds 5% above, 5% below, and at MLSS. A repeated trial at the MLSS was conducted during participants’ final visits. Participants were encouraged to run until volitional exhaustion up until a maximum duration of 45 min (excluding warm-up); however, data collected past 30 minutes were not included in the present study.

### Equipment and Measurements

#### Cardiorespiratory Measurements

All running tests were conducted on a treadmill (Desmo Pro Evo, Woodway USA Inc., Waukesha, WI, USA), with the treadmill incline set to 1% grade (Jones & Doust, 1996). Adjustments to treadmill speed could be made in 0.1 mph increments (i.e., ∼0.04 m·s^−1^). During all testing sessions, RR-interval data were collected using a Polar H10 chest strap (Polar Electro Oy, Kempele, Finland) with a sampling rate of 1000Hz, and recorded via Bluetooth using a Polar Vantage V (Polar Electro Oy, Kempele, Finland) watch. Ventilatory and gas exchange variables were collected using a mixing chamber (COSMED, Rome, Italy), facemask (7450 Series V2, Hans-Rudolph, Shawnee, KS, USA) connected to a 2-way non-rebreathing valve (Hans-Rudolph) and gas collection hose, and analyzed using a metabolic cart (Quark CPET, COSMED). The metabolic cart system was calibrated prior to each visit and 10-s averages of gas exchange data were used for analysis.

#### Blood Lactate Measurements

All [BLa] data were collected using capillary blood drawn from the fingertip and analyzed for [BLa] using the Biosen C-Line (EKF Diagnostics, Cardiff, Wales; n=6) or Lactate Plus (Nova Biomedical, Waltham, MA, USA; n=13) lactate analyzer. Runners straddled the treadmill (∼60-75s) during [BLa] measurements at 10 and 30 min (or task failure if < 30 min).

#### Data Processing

The interbeat RR-interval data output was imported into Kubios HRV Premium Software version 3.5.0 (Kubios Oy, Kuopio, Finland) for preprocessing with RR-interval detrending set to “smoothness priors” (Lambda = 500), automatic noise detection set to “medium”, and beat correction threshold set to “automatic.” RR-interval data files with noise or beat corrections that exceeded 3% were excluded from subsequent analysis (Rogers, Giles, Draper, et al., 2021b).

Monofractal DFAα1 was calculated using a linear fit of the double log RMS fluctuation data in relation to the different window sizes, with window width set to 4 :: n :: 16 (Peng et al., 1995). A modified Python script (Pickus, 2017) calculated DFAα1 from the processed RR-interval data files using 2 min of RR-interval data (Chen et al., 2002; Hautala et al., 2003) and re-calculated DFAα1 every 10 s. For example, the DFAα1 value corresponding to minute 8 in Figure 1 was calculated using RR-interval data collected from minutes 7 to 9 of the treadmill running protocols. This procedure was performed for all running trials.

**Figure 1.**
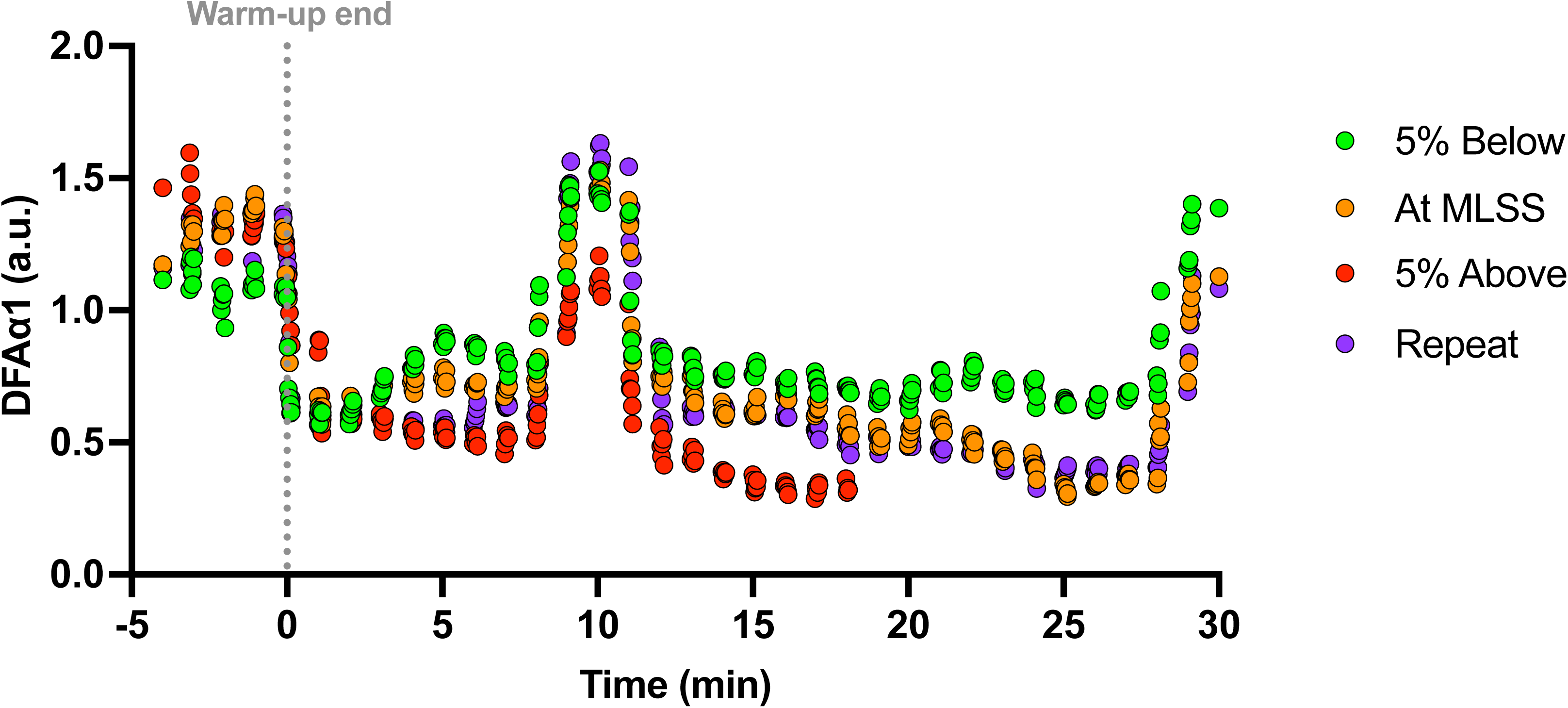
Example DFAα1 signal from one participant during the four constant-speed trials near the maximal lactate steady state (MLSS). Data are shown during 5 minutes of constant-speed running at 1.9 m·s^-1^ (i.e., warm-up) followed by constant-speed treadmill running for 30 minutes 5% below MLSS (3.4 m·s^-1^), at MLSS (3.6 m·s^-1^), and during a repeat trial at MLSS and ∼20 minutes of constant speed running 5% above MLSS (3.8 m·s^-1^). The participant straddled the treadmill (∼60-75s) during [BLa] measurements at 10 min and again at 30 min (or ∼20 min during their running trial 5% above MLSS).

^V̇^ O_2_max was identified as the highest 30s average ^V̇^ O_2_ achieved during the incremental test. The GET was identified as the ^V̇^ O_2_ associated with a disproportionate increase in the rate of carbon dioxide production (^V̇^ CO_2_) and minute ventilation (^V̇^ _E_) relative to the increase in ^V̇^ O_2_ (Beaver et al., 1986). The RCP was identified as the ^V̇^ O_2_ associated with a second disproportionate increase in ^V̇^ _E_ as well as a disproportionate increase in ^V̇^ _E_/^V̇^ CO_2_ relative to the increase in ^V̇^ O_2_ (Beaver et al., 1986). GET and RCP were determined by the primary researcher and two independent researchers prior to reaching a consensus.

#### HRV Threshold Determination

The following procedure was performed to identify the ^V̇^ O_2_ associated with DFAα1 values of 0.75 (i.e., HRV_GET_) and 0.50 (i.e., HRV_RCP_). First, the DFAα1 time-series data were time-aligned with ^V̇^ O_2_ measurements during the incremental treadmill running test.

Subsequently, linear regression was performed on a subset of the ^V̇^ O_2_-DFAα1 data starting from values near 1.0 to approximately 0.5, or below if values continued linearly, based upon previously established methodology (Rogers, Giles, Draper, Hoos, et al., 2021; Rogers, Giles, Draper, et al., 2021a). Using this linear equation, the ^V̇^ O_2_ values associated with a DFAα1 of 0.75 and 0.5 were identified as the HRV_GET_ and HRV_RCP_, respectively.

#### Constant Speed Treadmill Running

DFAα1 time-series data were averaged between min 7-8, 17-18, and 27-28 (or 1-min prior to the final minute before task failure) and used to represent the 10-min, 20-min, and 30-min time points during constant-speed running trials, respectively, to avoid any effect of the temporary stoppage in running to collect [BLa] on subsequent DFAα1 analysis. For example, the DFAα1 values corresponding to the 10^th^ minute represents the average DFAα1 measurement calculated between minute 7 and 8, which consists of RR-interval data collected from minutes 6 to 9 during the constant-speed treadmill running protocols (Figure 1). In addition, the ‘expected’ DFAα1 values during the constant-speed treadmill tests were calculated and used to represent the expected DFAα1 response during constant-speed treadmill running tests at the MLSS. This value was calculated as the DFAα1 value corresponding to the RCP using the linear ^V̇^ O_2_-DFAα1 regression from the incremental exercise test, and only calculated for participants who completed both incremental and constants-speed testing.

### Statistics

Paired Student’s *t* tests were used to compare pairs of variables. Effect sizes were calculated using Cohen’s d for paired variables (Cohen, 1992). Level of correlation was assessed using Pearson’s correlation coefficients. Agreement was assessed by Bland-Altman analyses (95% limits of agreement) and two-way mixed effects, absolute agreement, single rater intraclass correlation models, wherein reliability was interpreted as poor (ICC < 0.5), moderate (0.5 ≤ ICC < 0.75), good (0.75 ≤ ICC < 0.9), or excellent (ICC ≥ 0.9) (Koo & Li, 2016). Two-way repeated measures ANOVAs were used to assess the effects of intensity and duration on DFAα1 during constant-speed running trials near MLSS, and to test the effects of trial and duration between repeat trials at MLSS. Data are presented as mean [standard deviation (SD)]. Statistical significance was set at an α level of < 0.05. Statistical analyses were performed using Statistical Package for the Social Sciences (SPSS, version 26, IBM, Armonk, NY, USA), with data visualization performed using Prism (version 10.1.0 for macOS, GraphPad Software, San Diego, California USA).

## Results

### Data Inclusion

Data sets from participants exercise tests with RR-interval artifact percentages > 3%, in the RR-interval section of interest (Rogers, Giles, Draper, et al., 2021b), were removed from subsequent analysis, including two of 19 ramp-incremental tests (n=17); one of 12 constant-speed running tests (n=11); and two of 12 repeat trial tests (n=10).

### Participants

Table 1 displays participant characteristics, incremental exercise testing results, and HRV-derived thresholds. All incremental tests were maximal, and the duration of the incremental test portion of the SRS protocol was 12.5 [1.8] min. All twelve runners completed at least 30 min of treadmill running 5% below MLSS, at MLSS, and during the repeat trial at MLSS; however, four runners included in the final analysis were unable to complete 30 min of running 5% above MLSS.

### HRV Threshold Determination

Figure 2 displays the individual ^V̇^ O_2_-DFAα1 responses during incremental treadmill running and the linear regression equations used to derive HRV_GET_ and HRV_RCP._ The HRV_GET_ (42.3 [4.6] mL·kg^−1^·min^−1^) was strongly associated and had moderate agreement with the GET (38.7 [3.4] mL·kg^−1^·min^−1^; r=0.80, p<0.001; ICC=0.710, 95% C.I.=–0.193-0.916; Figure 3), but overestimated the GET by 3.6 mL·kg^−1^·min^−1^ (LOA: –9.1 to 1.9 mL·kg^−1^·min^−1^; p<0.001; d=1.28; Figure 3). The HRV_RCP_ (52.0 [8.0] mL·kg^−1^·min^−1^) was strongly associated and had moderate agreement with the RCP (48.6 [4.4] mL·kg^−1^·min^−1^; r=0.77, p<0.001; ICC=0.730, 95% C.I.=0.227-0.904; Figure 3), but overestimated the RCP by 3.5 mL·kg^−1^·min^−1^ (LOA: –14.1 to 7.2 mL·kg^−1^·min^−1^; p=0.019; d=0.63; Figure 3).

**Figure 2.**
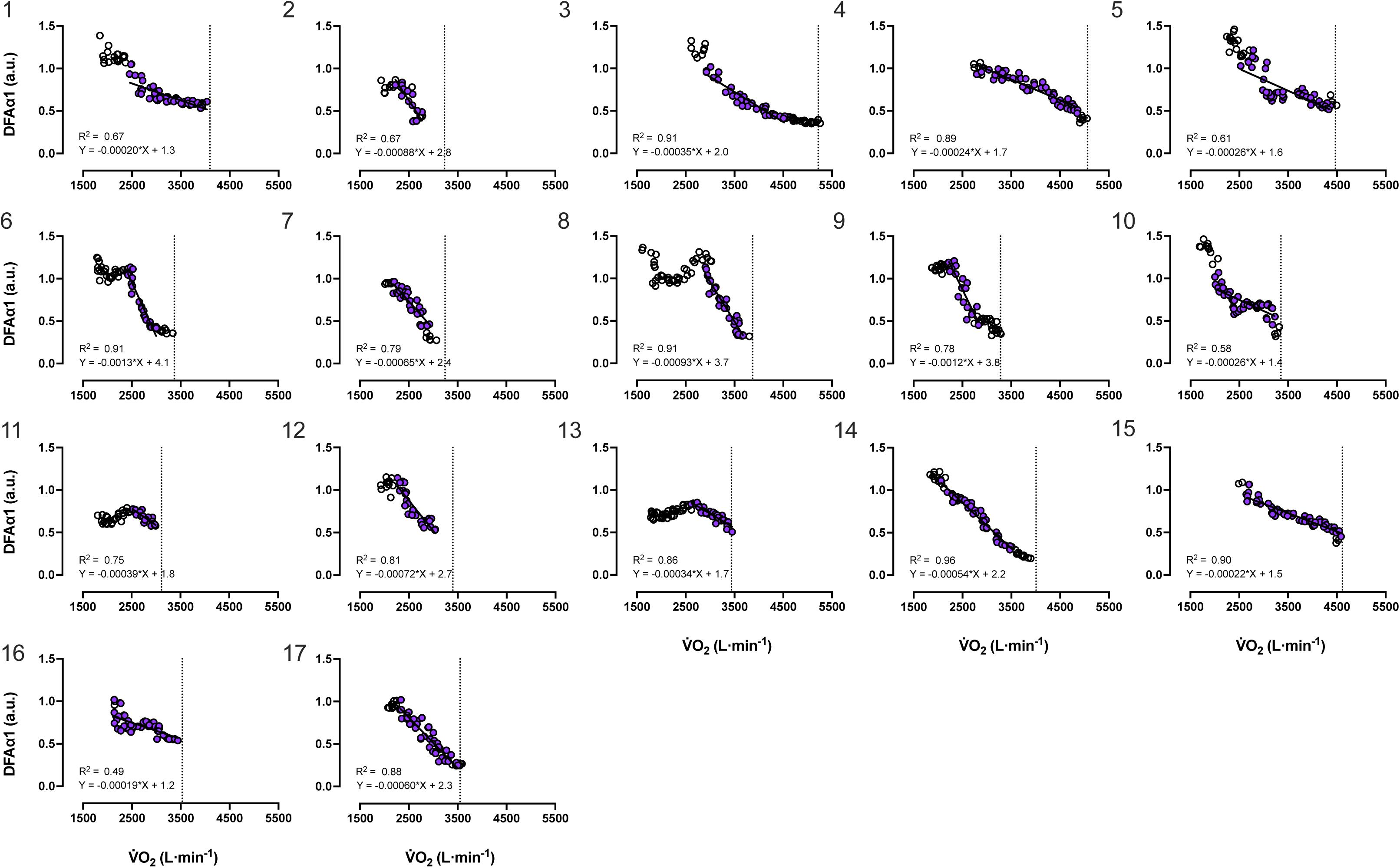
^V̇^ O_2_-DFAα1 responses for subjects 1 – 17 during incremental treadmill running. Purple circles indicate the DFAα1 values used to identify the HRV_GET_ and HRV_RCP_. Vertical dotted line indicates ^V̇^ O_2_max. Solid black line indicates trendline. DFAα1 values calculated using RR-interval data with artifacts >3% were removed from the visual (n = 17).

**Figure 3.**
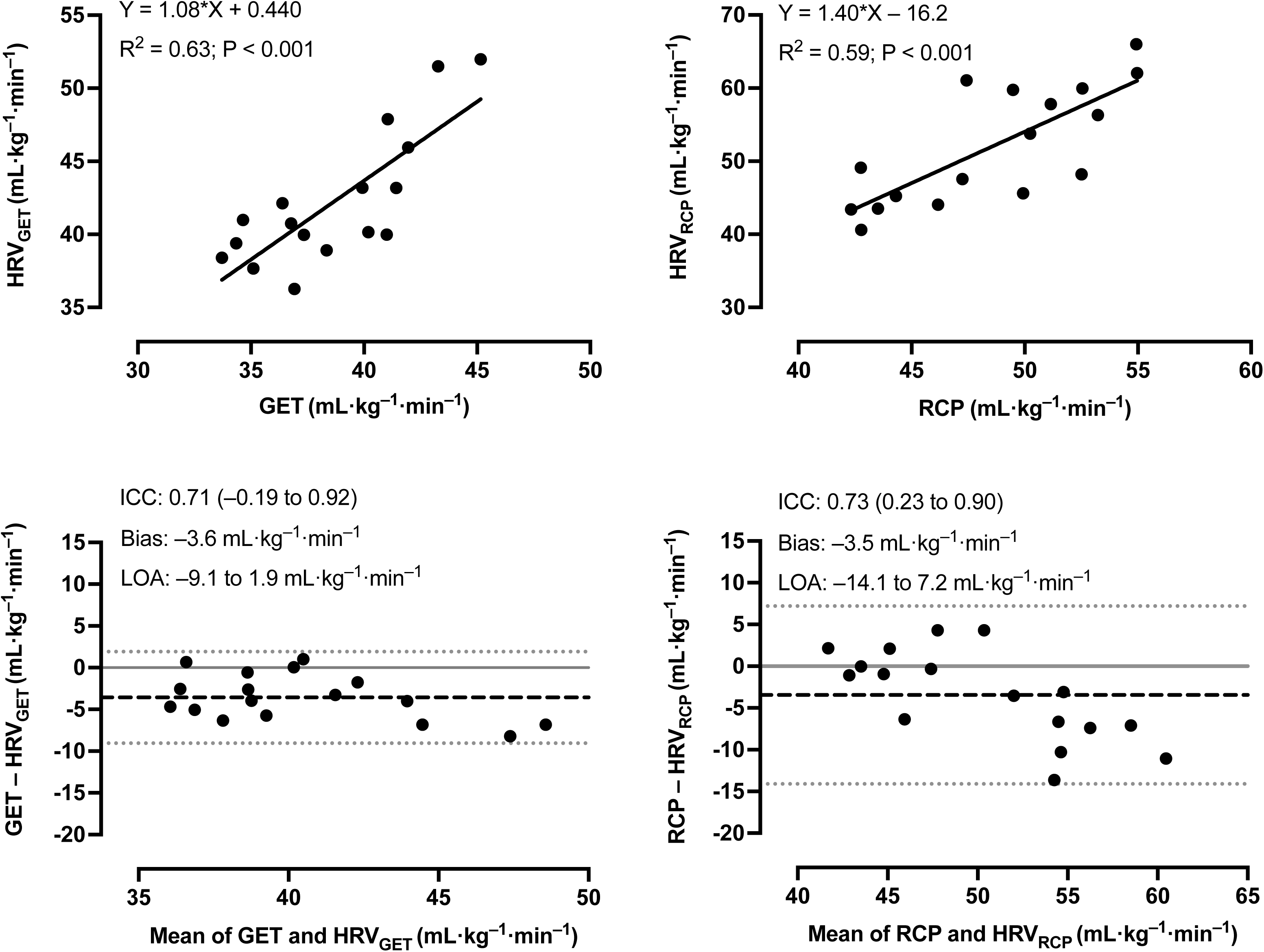
Panel A and B show the relationships between the GET and the HRV_GET_ and between the RCP and HRV_RCP_ derived from the incremental treadmill running tests. Panels C and D show Bland Altman plots corresponding to the data in Panels A and B, respectively. Dashed lines represent bias and dotted lines represent 95% limits of agreement. n=17 for all panels.

### Constant-Speed Running Near the MLSS

Figure 4 displays the DFAα1 responses at 10-, 20-, and 30-min (or task failure if < 30 min) during constant-speed running trials performed near MLSS. Mean [SD] DFAα1 measurements at 10-, 20-, and 30-min were 0.78 [0.26], 0.73 [0.23], and 0.62 [0.11] 5% below MLSS; 0.72 [0.12], 0.64 [0.13], and 0.50 [0.15] at the MLSS; and 0.66 [0.13], 0.48 [0.13] and 0.43 [0.09] 5% above MLSS. No significant intensity x duration interaction effect was detected for DFAα1 (p=0.207); however, there were main effects of intensity (p=0.019) and duration (p=0.001), with differences in DFAα1 measurements 5% below compared to 5% above MLSS (p=0.048) and at MLSS compared to 5% above MLSS (p=0.004), but not 5% below MLSS compared to at MLSS and differences between all time points (p <0.05 for all post hoc comparisons; Figure 4).

**Figure 4.**
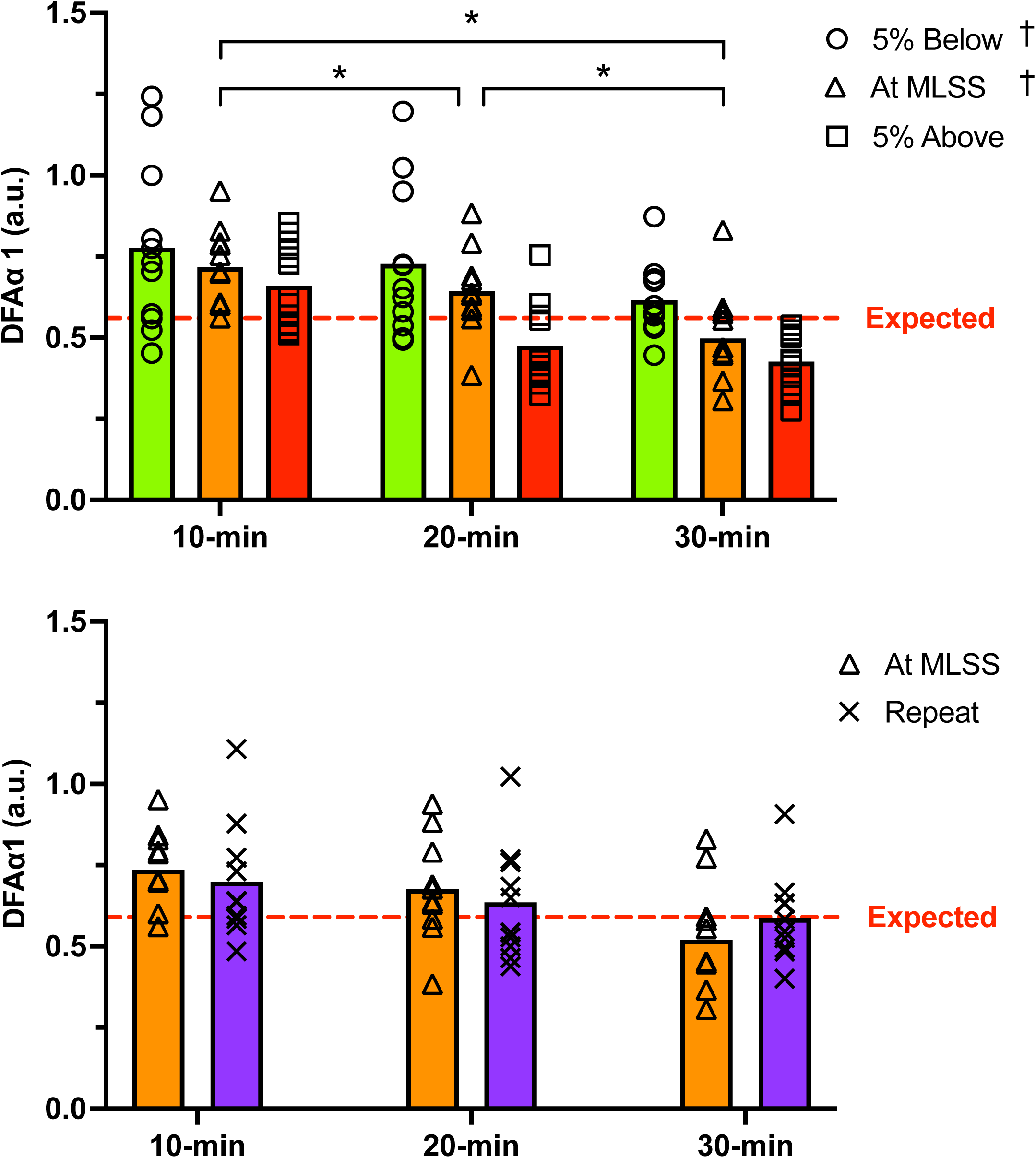
A) Comparison between DFAα1 measurements at 10-, 20-, and 30-min (or task failure if < 30 min) during constant-speed running trials performed 5% below (green), at (orange), and 5% above the maximal lactate steady state (MLSS) (red), and B) between repeat running trials at MLSS (purple). Dotted red line indicate the average DFAα1 corresponding to the RCP from the incremental running test, or the “expected” DFAα1 during constant-speed treadmill running. Asterisk (*) indicates significant difference between pairs of durations. Obelisk (†) indicates significant difference between the denoted intensity compared to 5% above the MLSS. Error bars represent one SD. n=11 and 10 for panels A and B, respectively.

The 10-, 20-, and 30-min DFAα1 measurements during repeat trials at MLSS are presented in Figure 4. A significant trial x duration interaction effect was detected for DFAα1 between MLSS trials (p=0.022) as well as a main effect of duration (p=0.014). No main effect of trial was detected (p=0.926). Pairwise comparisons did not reveal a significant difference in DFAα1 between trial or duration pairings (p>0.05 for all post hoc comparisons; Figure 4).

## Discussion

In support of recent investigations (Mateo-March et al., 2023; Rogers, Giles, Draper, Hoos, et al., 2021; Rogers, Giles, Draper, et al., 2021a; Schaffarczyk et al., 2023), the present study detected strong correlations and moderate agreement between the HRV_GET_ and GET and between the HRV_RCP_ and RCP; however, the HRV method overestimated both the GET and RCP and produced relatively wide LOA between HRV_RCP_ and RCP measures. Further, in contrast to our hypothesis, 30-min of constant-speed treadmill running at the MLSS (i.e., the upper boundary of heavy intensity domain) yielded a wide range of DFAα1 values (i.e., 0.31 to 0.95), indicative of exercise intensities spanning the three intensity domains if the threshold values of 0.75 and 0.5 were considered (Mateo-March et al., 2023; Rogers, Giles, Draper, et al., 2021a).

Overall, our results suggest that the HRV method may provide inaccurate estimations of exercise thresholds and that the intensity-dependent DFAα1 response during incremental running is not equivalent to the intensity-dependent DFAα1 response during constant-speed running. Despite this dissociation, the DFAα1 response during constant speed running was intensity-and duration-dependent and repeatable, suggesting it may provide a metric that can quantify systemic perturbations and characterize physiological shifts imposed by intense, continuous exercise.

In contrast to our hypothesis, fixed DFAα1 values did not accurately identify exercise thresholds. On average, the HRV_GET_ overestimated the GET, but in alignment with previous investigations (Mateo-March et al., 2023; Rogers, Giles, Draper, Hoos, et al., 2021; Schaffarczyk et al., 2023), the two measurements were strongly correlated and had comparable LOA. Similarly, the HRV_RCP_ overestimated the RCP, and the two measures were strongly associated; but in contrast to the HRV_GET_, the LOA between the HRV_RCP_ and RCP were wide. Wide LOA between evaluations of the RCP and HRV_RCP_ have previously been found (Mateo-March et al., 2023; Rogers, Giles, Draper, et al., 2021a). Of note, it appears that the agreement between the HRV_RCP_ and RCP measurements was influenced by runner fitness level, as RCP measurements, which are directly associated with running performance capacity (Bassett & Howley, 2000), were higher in runners with larger biases between the HRV_RCP_ and RCP (Table 1; Figure 3). Accordingly, our results suggest that using fixed DFAa1 values 0.75 and 0.50 to identify ventilatory threshold during incremental running may produce variable and systemic overestimations of exercising intensity, particularly when using this method to detect the RCP.

The intensity-dependent response of DFAα1 during incremental exercise has been repeatedly reported (Mateo-March et al., 2023; Rogers, Giles, Draper, Hoos, et al., 2021; Rogers, Giles, Draper, et al., 2021a; Schaffarczyk et al., 2023). Generally, this association is described as an inverse relationship with DFAa1 values starting between ∼1.0-1.5 at moderate intensities, decreasing to ∼0.75 as intensity transitions from the moderate to the heavy domain, and continuing to drop past 0.5 as intensity transitions from the heavy to the severe domain. Along with non-neural factors that adjust during exercise, these responses are attributable to parasympathetic nervous system withdrawal and sympathetic nervous system activation as exercise intensity increases (Persson, 1996; Shaffer & Ginsberg, 2017; White & Raven, 2014). In support of this well documented intensity dependent DFAα1 response, we found similar—albeit, highly variable—DFAα1 responses during incremental running (Figure 2). For example, the DFAα1 range and ^V̇^ O_2_-DFAα1 slopes (calculated using DFAα1 values between ∼1.0 and 0.5) were highly variable, with some runners DFAα1 values never exceeding 1.0 or falling below 0.5 (Figure 2). There also appeared to be a relationship between fitness level and the capacity for DFAα1 to fluctuate, as fitter runners (i.e., higher ^V̇^ O_2_max) seemed to demonstrate a more dynamic DFAα1 response including larger ranges in DFAα1 (i.e., highest vs. lowest values) and more gradual declines in DFAα1 compared to less fit runners (i.e., lower ^V̇^ O_2_max) (Figure 2). Accordingly, our results support an intensity-dependent DFAα1 response during incremental treadmill running that is related to exercising thresholds and DFAα1 values of 0.75 and 0.5.

However, rather than indicating exercising thresholds, it appears that the DFAα1 response may provide an indication of the physiological perturbation caused by incremental exercise, which— although highly related to exercise intensity—may not necessarily provide an accurate indication of exercise thresholds. In alignment with this “physiological perturbation” hypothesis, repeated incremental running has previously been found to shift the agreement between ventilatory threshold and DFAα1 derived thresholds (Van Hooren et al., 2023). In the current investigation, the HRV_RCP_ seemed to be overestimated by a larger degree in fitter runners (i.e., higher relative RCP; Figure 3), suggesting that fitter runners may have a greater capacity to resist physiological perturbation from incremental running, while the narrow DFAα1 response in less fit runners (i.e., lower ^V̇^ O_2_max; Figure 2) may suggest that the physiological demand of the incremental running test was relatively high.

In further support of this speculation, the findings from our constant-speed running trials near MLSS provide evidence that both exercise intensity and duration affect the DFAα1 response (Figure 4). These responses were repeatable and independent of exercise intensity relative to the MLSS (Figure 4). Indeed, using the fixed DFAα1 framework, DFAα1 measurements after 10 minutes of constant-speed running 5% above the MLSS, were indicative of exercise within the heavy intensity domain (i.e., averaging 0.66), with some runners displaying HRV correlation values indicative of exercise in the moderate intensity domain (i.e., > 0.75; Figure 4). Although constant-speed running at MLSS did produce DFAα1 responses within the expected range for heavy intensity exercise (i.e., between 0.75 and 0.5), some runners’ DFAα1 values remained above this range after 30 min of exercise at the MLSS despite DFAα1 clearly responding to exercise duration (Figure 4). Accordingly, during constant-intensity exercise, it does not appear that DFAα1 provides an indication of exercising intensity domain and may instead indicate the accumulation of exercise fatigue or overall physiological demand of a given task. Previous investigations have found similar effects of duration on DFAα1, with an attenuation in the DFAα1 signal during a 10-km race in slower runners (Gronwald, Hoos, et al., 2020); a reduction in DFAα1 measures following an ultramarathon (Rogers, Mourot, et al., 2021); and a continued decrease in DFAa1 measures over the course of a marathon (Gronwald et al., 2021). The results from our investigation also suggest that these responses are repeatable over a given duration and intensity (Table 2; Figure 4), and support previous suggestions (Gronwald et al., 2021; Rogers & Gronwald, 2022) that DFAα1 represents an indication of the systemic demand of exercise.

### Experimental considerations

Previous investigations suggest that maximal exercise immediately preceding an incremental test can affect the agreement between ventilatory and DFAα1 threshold (Van Hooren et al., 2023). Accordingly, the 18-minute MOD step performed immediately before the incremental test may have affected the agreement between ventilatory and DFAα1 measures in the current investigation; however, due to the overestimation of ventilatory thresholds, it is possible that moderate intensity exercise prior to incremental exercise may attenuate the DFAα1 response. With consideration to the highly variable ^V̇^ O_2_-DFAα1 slopes observed during incremental exercise, slight changes to the starting points (i.e., DFAα1 values near 1.0) and finishing points (i.e., DFAα1 values near 0.5) included in the linear regression equation can dramatically affect HRV threshold estimations; therefore, the effect of model fitting and model type (e.g., linear vs. polynomial) on HRV threshold determination should be investigated further. It should also be noted that detecting GET and RCP is subject to inter-rater biases, and although a consensus between three researchers was made prior to determining the GET and RCP, inaccurate assessments may have influenced the agreement ventilatory and HRV thresholds. Finally, our results should be considered with respect to our method of intensity domain demarcation as the MLSS may indeed represent the upper boundary of heavy intensity exercise, while metrics such as critical speed or critical power may provide a better representation of the MMSS boundary itself (Nixon et al., 2021).

## Conclusion

The results from this investigation provide additional evidence supporting the strong associations between the DFAα1 threshold detection methodology with the GET and RCP but questions the utility of this method to accurately detect exercise thresholds in practice. Our results suggest that incremental exercise does not produce equivalent HRV correlation profiles as constant-speed treadmill running near MLSS and that DFAα1 should not be used to indicate exercise intensity domain during constant speed treadmill running. DFAα1 measures at MLSS were repeatable and affected by both duration and intensity. In conclusion, our study suggests that DFAα1 offers a dimensionless index of the complex regulation of human activity during exercise that may indicate fatigue accumulation and physiological perturbation from prolonged exercise stress. This metric may provide utility to characterize the physiological shifts inflicted by high-intensity continuous exercise to profile a runner’s performance capacity.

## Data Availability

All data produced in the present work are contained in the manuscript

## Acknowledgments

This investigation was supported by an operating grant from the Natural Sciences and Engineering Research Council of Canada (NSERC; grant number RGPIN-2018-06424) and start-up funding from the Faculty of Kinesiology (University of Calgary) received by MJM. CVR was funded by NSERC, the NSERC CREATE Wearable Technology and Collaboration (We-TRAC) Training Program, an Alberta Innovates Graduate Student Scholarship for Data-Enabled Innovation, and an Alberta Graduate Excellence Scholarship. The authors would like to acknowledge the contributions of all participants, students, faculty, and staff, who assisted and made this investigation possible.

